# Machine Learning-Driven Identification of Serotype-Independent Pneumococcal Vaccine Candidates using samples from Human Infection Challenge Studies

**DOI:** 10.1101/2025.09.12.25335625

**Authors:** Katerina S. Cheliotis, Patricia Gonzalez-Dias, Esther L. German, André N. A. Gonçalves, Elena Mitsi, Elissavet Nikolaou, Sherin Pojar, Eliane N. Miyaji, Rafaella Tostes, Jesús Reiné, Andrea M. Collins, Helder I. Nakaya, Stephen B. Gordon, Ying-Jie Lu, Shaun H. Pennington, Andrew J. Pollard, Richard Malley, Simon P. Jochems, Britta Urban, Carla Solórzano, Daniela M. Ferreira

## Abstract

Identifying conserved, immunogenic proteins that confer protection against *Streptococcus pneumoniae* colonisation could enable development of serotype-independent vaccines.

We analysed baseline samples from 86 healthy adults experimentally challenged with pneumococcal serotypes 6B or 15B to investigate whether immune responses to 75 universally expressed pneumococcal proteins associated with protection against colonisation. We measured serum IgG using a novel Luminex assay and cytokine responses from peripheral blood mononuclear cells following antigen stimulation.

No individual IgG or cytokine marker correlated significantly with protection in univariate analysis. However, machine learning identified IgG responses to PdB, SP1069, and SP0899 as predictive of protection. MCP-1 responses to SP1069 and SP0899, and IL-17 production in response to SP0648-3 also correlated with protection. Elevated baseline IFN-γ, RANTES, and anti-protein IgG correlated with lower colonisation density.

We highlight SP1069 and SP0899 as potential serotype-independent vaccine candidates and demonstrate the utility of machine learning to identify immune correlates of protection.

**Graphical Abstract:** 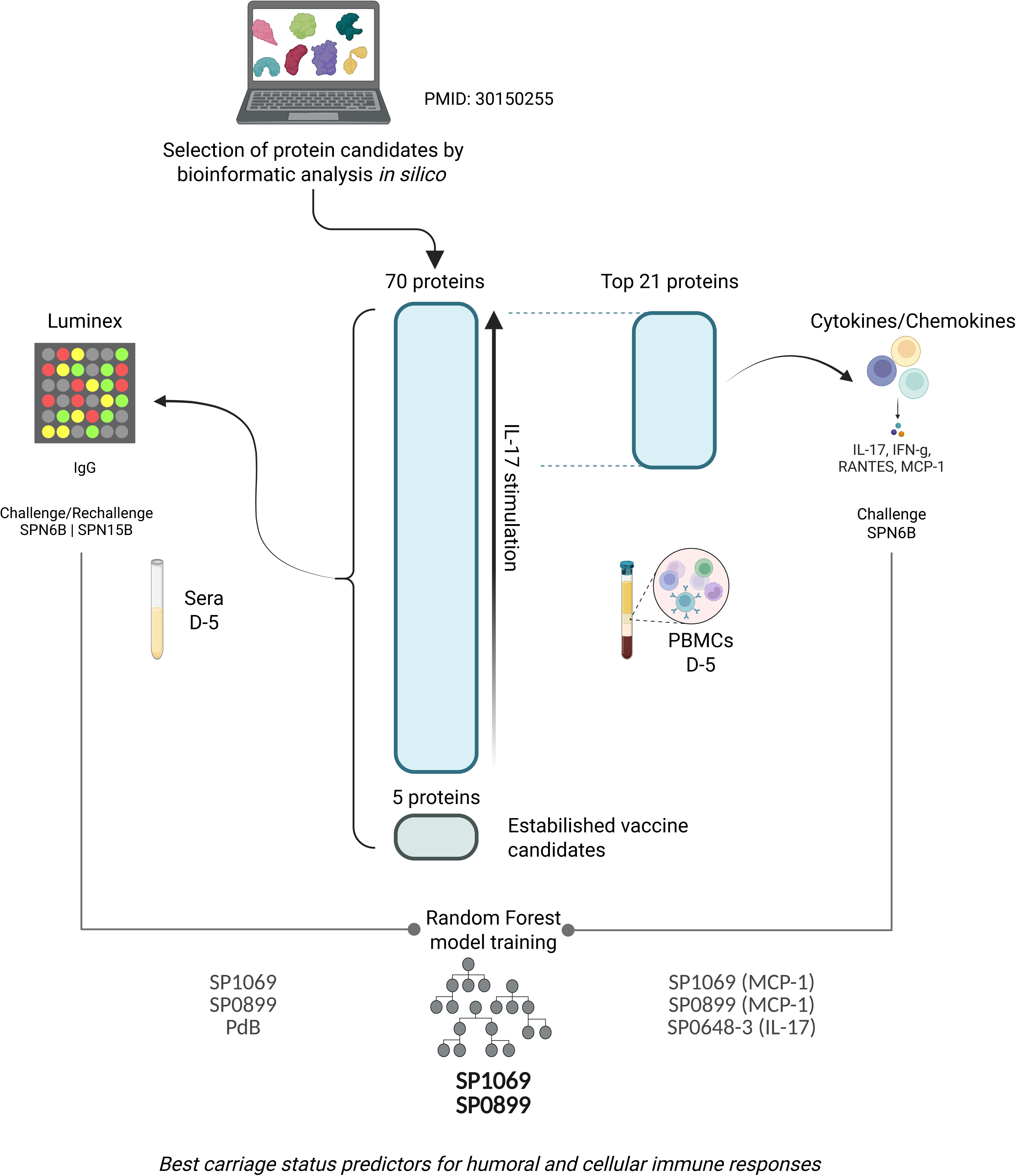

## Introduction

Pneumonia is the leading infectious cause of death in infants worldwide, with more than 800,000 annual deaths among children under 5 years of age globally ^1–3^. The most common aetiological agent of pneumonia is the bacterium *Streptococcus pneumoniae.* Nasopharyngeal colonisation with pneumococcus, as well as being a prerequisite for invasive disease, is a source of transmission in the community ^4^.

The pneumococcal conjugate vaccine (PCV) generates population herd protection providing substantial cost-benefit of vaccination ^5^. Vaccine-induced humoral responses to the pneumococcal capsule polysaccharide protect against pneumococcal carriage acquisition and disease caused by vaccine specific serotypes ^6–10^. However, current vaccines – pneumococcal polysaccharide vaccine (PPV23) and PCVs – offer protection against a limited number of the >100 pneumococcal serotypes ^11^ leading to serotype replacement of non-vaccine types ^12,13^. In addition, despite being included in PCVs, serotype 3 continues to circulate and cause a high disease burden particularly in older adults ^14–16^. PCVs are also complex and costly to produce, which can present a barrier to vaccine access in resource-limited countries, particularly those graduating from GAVI support ^17^.

Pneumococcal protein antigens that are universally expressed across various serotypes may induce both humoral and cellular immune responses, offering broad protection. These antigens could potentially be introduced as stand-alone vaccines or be included as conjugate proteins in current polysaccharide-based vaccines ^18^.

Currently the regulatory approval pathway for next-generation pneumococcal vaccines in infants relies on the non-inferiority of IgG levels to capsular polysaccharide using a pre-defined threshold of 0.35ug/ml. In adults demonstration of non-inferior or superior opsonophagocytic antibody responses to each serotype compared to existing licensed vaccines is required^19^. Therefore, the regulatory pathway for capsular polysaccharide independent vaccines is not defined. The identification of serotype-independent cellular and humoral correlates of protection against pneumococcal infection could assist with this framework and help to accelerate development of serotype-independent vaccines.

Systems-biology and artificial intelligence can be used to significantly enhance our understanding of immunity and aid in the identification of correlates of protection. Embedding the understanding of individual components and their interactions, beyond canonical antibody binding and neutralisation, may accelerate the design of more successful and long-lasting vaccines ^20^.

In this study, we leveraged a controlled human infection model (CHIM) to investigate immune responses to pneumococcal proteins that are conserved across serotypes, aiming to identify correlates of serotype-independent protection and to understand their impact on colonisation dynamics in the nasopharynx. To enable high-throughput assessment of protein-specific IgG responses, we established a Luminex assay targeting 75 pneumococcal protein antigens. Given the well-established role of IL-17 in mediating protection against *S. pneumoniae* in murine models, we also evaluated IL-17-driven cellular responses in protein-stimulated peripheral blood mononuclear cells (PBMCs). In parallel, we employed a multiplex cytokine assay to profile broader cytokine responses, with MCP-1 (CCL2), RANTES (CCL5), and IFN-γ selected for more detailed analysis.

## Methods

### Study Design

The overall study design is shown in Figure 1. Briefly, at baseline (day −4 and day −5), participants were assessed before intranasal inoculation (day 0) with *S. pneumoniae* (Spn). Carriage acquisition was monitored until day 14 for Spn15B and day 29 for Spn6B.

**Figure 1.**
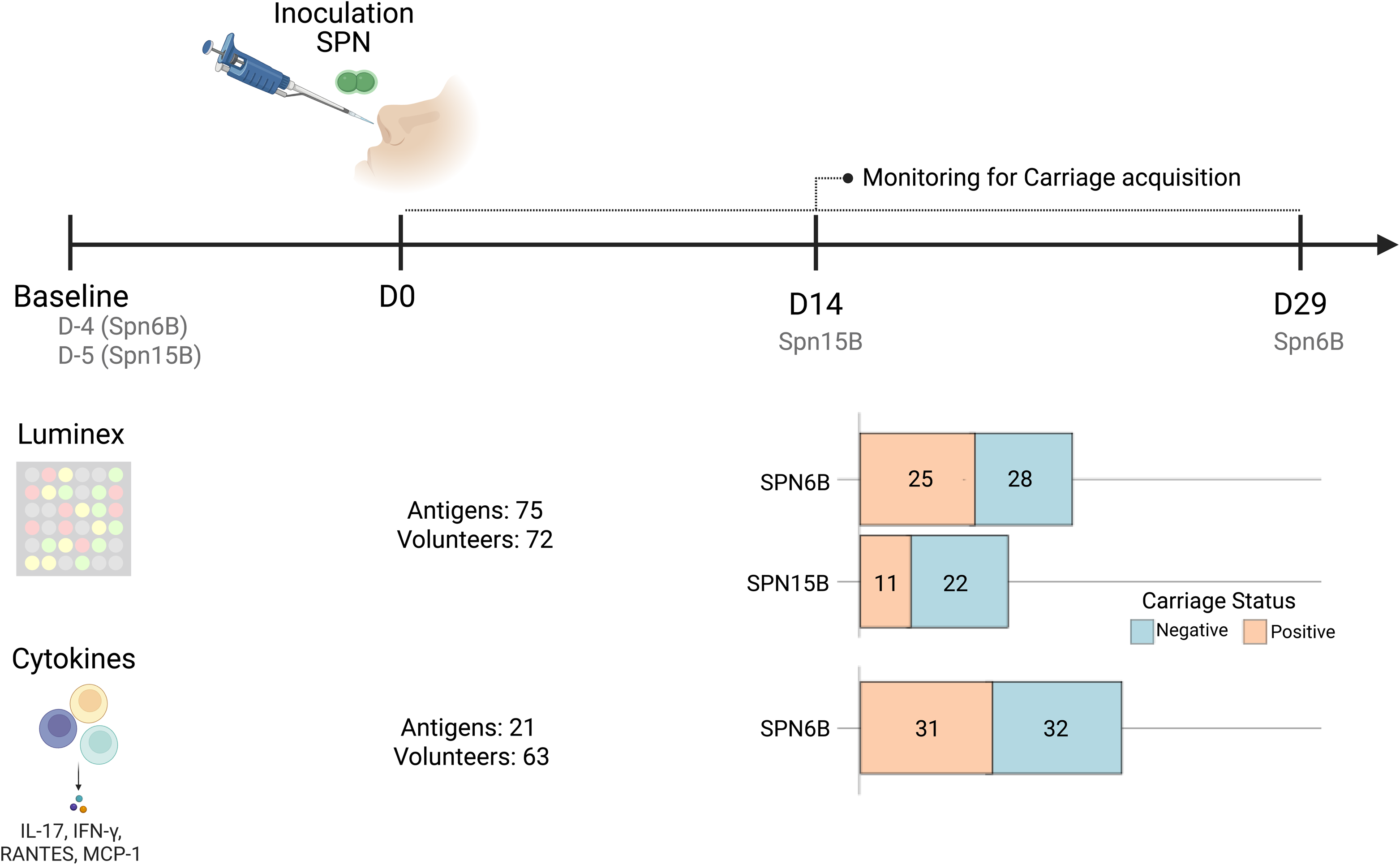
Study design. Participant serum was collected at ‘Baseline’ (4 days for Spn6B and 5 days for Spn15B) prior to experimental inoculation with pneumococcus (Spn) at day 0 (D0), and carriage acquisition assessed until day 29 post-challenge (days 2, 6, 9, 14 and 29 post-challenge with Spn6B; days 2, 7 and 14 post-challenge with Spn15B). Antibody responses were evaluated using the Luminex assay. Cytokine/chemokine responses (IL-17A, IFN-γ, RANTES (CCL-5) and MCP-1 (CCL-2)) following the stimulation of PBMCs with pneumococcal protein antigens were evaluated by ELISA. The number of antigens and the number of participants per carriage status are shown. Bar plots show the number of participants that were colonised with pneumococcus (orange) and those that were protected against carriage acquisition (blue).

Immune responses were evaluated using Luminex assays, which measured IgG antibody responses to 75 pneumococcal antigens in 72 volunteers, and cytokine profiling, which detected 21 immune markers (including IL-17, IFNγ, RANTES, and MCP-1) in 63 volunteers.

Carriage acquisition results are presented as bar graphs, depicting the number of individuals testing positive or negative for Spn6B and Spn15B at different time points. These results indicate the proportion of participants who acquired pneumococcal carriage following inoculation.

### Participant cohort inoculated with serotype 6B pneumococcus

Blood samples were taken from healthy non-smoking participants between the ages of 18 and 50 who were intranasally inoculated with 80,000 CFU in 0.1ml solution of *S. pneumoniae* serotype 6B (Spn6B) strain BHN418 per naris (full sequence available GenBank: ASHP00000000). Samples were taken prior to inoculation (baseline). Cohorts were recruited as part EudraCT (2014-004634-26), a study to evaluate the effect of live attenuated influenza vaccine (LAIV) on experimental human pneumococcal colonisation (EHPC) ^21,2221,22^. All participants gave written informed consent, with approval from the North West NHS Research Ethics Committee (14/NW/1460). Nasal wash samples were taken on days 2, 6, 9, 14 and 29 post-challenge with Spn6B. Colonisation was determined by classical microbiology and the presence of Spn6B in nasal wash samples taken at pre-defined time-points between inoculation and 29 days post-inoculation. No participants had previously received any pneumococcal vaccine.

### Participant cohort inoculated with serotype 15B pneumococcus

Healthy, non-smoking participants aged 18-50 years were intranasally inoculated with 80,000 CFU *S. pneumoniae* serotype 15B (Spn15B) per naris (clinical isolate 15B P1262. European Nucleotide Archive accession number: ERS2632437). Spn15B is included in the PPV23 formulation but not in the 13-valent PCV formulation. All participants gave informed written consent, ethical approval was obtained from the National Health Service Research Ethics Committee, Liverpool East (15/NW/0931). Serum samples were taken at baseline (pre-inoculation) (Trial identifier: ISRCTN 68323432; 20815). Colonisation was determined by classical microbiology and the presence of Spn15B in nasal wash samples taken at pre-defined time-points between inoculation and 14 days post-inoculation. Nasal wash samples were taken on days 2, 7 and 14 post-challenge with Spn15B. No participants had previously received any pneumococcal vaccine.

### Luminex assay

The proteins used in the assay are given in Supplementary Table 1. The sequences encoding the alpha-helical N-terminal domain of PspA1, PspC6, PspC9, were cloned using DNA from strain BHN418 (serotype 6B pneumococcus). The sequence encoding the N-terminal domain of PspA4 was cloned from strain 255/00 (serotype 14, GenBank accession number EF649969).

Recombinant proteins were produced at Butantan Institute laboratories as described previously^23^ including pneumolysin toxoid B (PdB), a pneumolysin mutant with a tryptophan-to-phenylalanine substitution at position 433 and 0.1% haemolytic activity of wild-type pneumolysin ^2425^. The remaining 70 proteins in the library were produced at Boston Children’s Hospital as described ^26^.

All 75 pneumococcal protein antigens were conjugated to magnetic microspheres at a concentration of 5µg per million microspheres using the xMAP Antibody Coupling Kit as per the manufacturer’s instructions. The assay was validated for the detection of anti-protein IgG as described previously ^27,28^. Full methods are described in detail in Supplementary Text 1. A stock of pooled sera taken 29 days post-challenge and diluted 1:50 in assay buffer was used as an internal positive control across all plates and assay buffer was used as a negative control to detect background median fluorescence intensity (MFI) values.

### Luminex data cleaning and statistical analysis

Raw Luminex data were cleaned prior to analysis through the following workflow. The coefficient of variation (CV) of replicates was calculated for all analytes. If any analyte had greater than 25% CV and both replicates had greater than 50 microspheres per well, the analyte was removed from analysis. If any paired samples had less than 50 microspheres of a given analyte in each well, the analyte was removed from analysis. The determinant of 50 microspheres is based on recommendations by the Luminex Corporation that a recovery of less than 50 microspheres renders a result unreliable as well as previous work showing that lower bead counts elicit higher CV values ^29^. All statistical analysis was carried out in RStudio (version 1.0.153). Statistical significance was determined via a Mann-Whitney or Wilcoxon test for unpaired and paired groups, respectively, followed by Benjamini Hochberg correction for multiple comparisons. For the machine learning analysis, any proteins with greater than 25% of values missing and any participants with more than 50% of values missing were removed from the analysis.

### Stimulation of human peripheral blood mononuclear cells (PBMCs) and cytokine/chemokine detection

Baseline PBMCs from healthy participants inoculated with Spn6B were thawed with 50 µg/ml DNAse I (MilliporeSigma) in prewarmed RPMI containing 10% FBS and washed once in media including DNAse I. 1×10^6^ PBMCs/ml from an initial cohort of 12 carriage-negative and 11 carriage-positive participants were stimulated with 8µg/ml of each protein from the complete library and 5µg/ml of heat-inactivated Spn6B for 7 days at 37°C under 5% CO_2_. The plates were then centrifuged to pellet the cells, after which the supernatants were collected to measure IL-17A concentration using human ELISA kits (Invitrogen). In parallel, supernatants from carriage-negative and carriage-positive participants from this initial cohort were combined and assessed for other cytokine and chemokine concentrations using a 30-plex magnetic human Luminex cytokine kit (Thermo Fisher Scientific) and acquired on an LX200. A log 2-fold change between carriage-negative and positive was calculated and a Wilcoxon paired test was used to identify cytokines/chemokines associated with a specific carriage status.

Twenty-one antigens from a 70-protein antigen library were selected for further analysis. The antigens were selected based on (i) their ability to induce high IL-17A responses in carriage-negative individuals (ii) negative correlation between high IL-17A levels and low pneumococcal density in participants who were experimentally colonised (iii) their ability to induce high expression of cytokines/chemokines at baseline prior to challenge (iv) available published data in which the antigens have shown protection against challenge after immunisation in pre-clinical models of infection.

PBMCs from an additional 22 carriage-negative and 21 carriage-positive participants were stimulated with the selected 21 antigens and cytokine/chemokine concentrations were measured to validate the results obtained with the initial cohort. Human ELISA kits were used to measure concentrations of IL-17A (Invitrogen), MCP-1 (CCL-2) (BD OptEIA^TM^), IFN-γ (BD OptEIA^TM^), and RANTES (CCL-5) (Biolegend) in the collected supernatants following the manufacturers recommendations. Non-stimulated (control) and protein-stimulated supernatants were diluted as follows: 1:2 for IL-17, 1:10 for RANTES, 1:200 for IFN-γ, and 1:3200 for MCP-1.

### ELISA data cleaning and statistical analysis

As detailed above, any proteins with greater than 25% of values missing and any participants with more than 50% of values missing were removed from the analysis. Statistical significance was determined via a Wilcoxon test for unpaired groups, followed by Benjamini Hochberg correction for multiple comparisons.

### Area under the curve (AUC) calculation

Area under the curve (AUC) was used to calculate overall colonising density of *S. pneumoniae* in the nasopharynx of experimentally colonised participants. AUC was calculated by trapezoidal rule from log 10 transformed density results at 2, 6, 16, 22, 27 and 36 days or after challenge on day 2, 7 or 14 with interpolation of missing data assuming a linear change in density between known values. The AUC of pneumococcal colonising density at each timepoint was derived and summarised using the above descriptive statistics and analysed using a generalised linear model with a single factor of treatment.

### AUC correlation analysis

A Spearman test was performed to evaluate the correlation between the IgG and cytokine/chemokine levels elicited in response to each protein and the AUC values obtained between days 2 to 14 for each carriage-positive participant using the cor.test function in the software R. The analysis was performed using antibody response and cellular response from 34 and 27 participants, respectively. Correlations with *p*-value ≤ 0.05 were considered significant.

### AUC threshold definition

Retrospectively, we calculated the AUC of nasopharyngeal Spn6B density between days 2 to 14 post-challenge in 128 experimentally colonised participants across our CHIM studies to define “Low”, “Mid” or “High” colonising density. For participants experimentally colonised with Spn15B, threshold AUC values were set using colonising density between days 2 to 14 from 9 participants. Threshold values differed for the two serotypes. The data distribution was used to split the groups for both Spn6B and Spn15B inoculated cohorts. The participants from different AUC groups were localised in different quartiles (Supplementary Figure 1).

### Machine learning analysis

Missing values in numerical variables were imputed using the “mice” R package ^30^. Numeric features were standardized using z-score normalization. The Random Forest algorithm, an ensemble machine learning method that constructs multiple decision trees using random feature subsets and aggregates their outputs to enhance predictive accuracy and minimize overfitting (28), implemented in the R package “randomForest”. The model was trained with mtry = 1 and ntree = 500, while other hyperparameters were set to their default values. This algorithm was employed to identify baseline protein-specific antibodies and cytokines/chemokines produced in response to PBMC stimulation with protein antigens that best predicted carriage status post-challenge. Unlike univariate analysis, which assesses each feature independently, machine learning techniques such as Random Forest can uncover complex, multivariate relationships among features, improving predictive accuracy.

Feature importance was assessed using the mean decrease in Gini index, a metric that quantifies how much a given feature contributes to reducing impurity (i.e., improving class separation) within the decision trees. A higher mean decrease in Gini index indicates a stronger contribution to classification accuracy.

To identify the most relevant features and optimize model performance, we applied an iterative feature selection process based on feature importance. Initially, we trained the Random Forest model using all 75 available features and evaluated their importance based on the mean decrease in Gini index. We then progressively removed the least informative features, retraining the model at each iteration and assessing its predictive performance. This process was repeated several times, testing progressively smaller feature sets (e.g., 50, 40, etc.), until the optimal configuration was identified, the smallest number of features yielding the best model performance, minimizing error, and reducing the risk of overfitting. Model evaluation was conducted using 10-fold cross-validation.

## Results

### Serum IgG levels to 75 different protein antigens at baseline did not associate with protection against experimental pneumococcal carriage acquisition of Spn6B or Spn15B

A Luminex-based multiplex assay using a library of 75 highly conserved pneumococcal proteins (Supplementary Table 1), was successfully optimised and validated ^28^. No associations between baseline levels of serum IgG against any of the protein antigens and protection against carriage acquisition remained statistically significant after applying the Benjamini-Hochberg correction for multiple testing (Figure 2).

**Figure 2.**
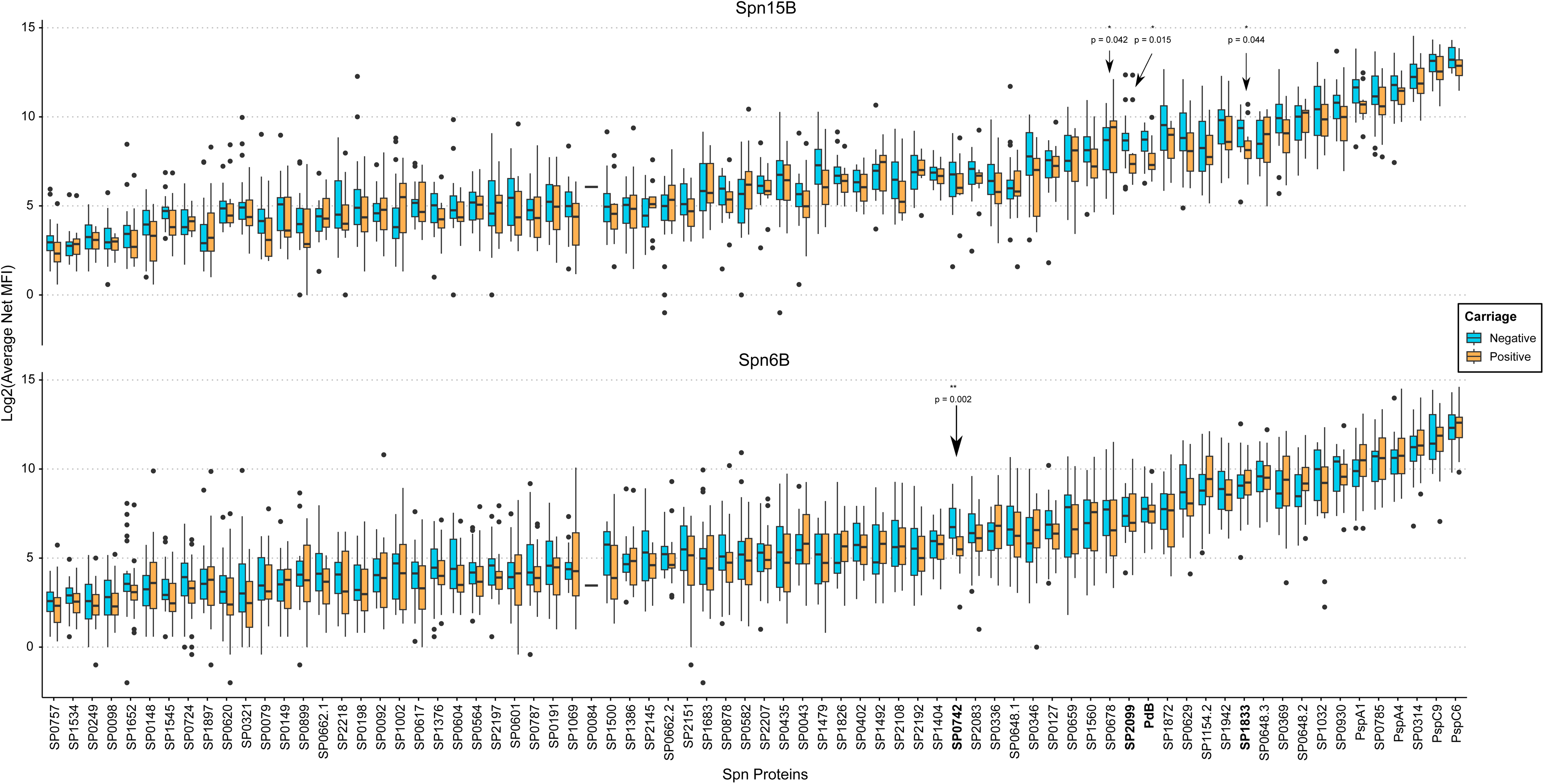
Levels of anti-protein IgG at baseline in participant cohorts subsequently challenged with Spn6B or 15B. Baseline levels of anti-protein serum IgG in healthy adults aged 18-50, measured as average net median fluorescence intensity (MFI). Bars indicate interquartile range. Spn15B carriage-negative n = 15, Spn15B carriage-positive n = 9; Spn6B carriage-negative n = 18, Spn6B carriage-positive n = 21. Responses with p-value ≤ 0.05 prior to Benjamini-Hochberg correction for multiple comparisons are shown; this significance was lost following correction.

### Screening of antigens using human PBMCs and cellular responses

A library of pneumococcal protein antigens was used to stimulate PBMCs collected from participants prior to experimental challenge with Spn6B. Due to the limited number of PBMCs obtained from each participant, we were not able to screen all protein antigens in all participants. IL-17A production has previously been described as a marker of protection against pneumococcal colonisation in mice^31,32^. IL-17A secretion following ex-vivo protein stimulation was observed for all proteins at 7 days of culture.

To identify other cellular markers that could associate with a protective or susceptible profile in the CHIM, we pooled supernatants from PBMC cultures following individual protein stimulation from carriage-negative and carriage-positive participants and compared levels produced of 30 cytokines and chemokines using 30-plex Luminex in these two groups (Supplementary Figure 2). MCP-1 (CCL-2) was selected as a marker of protection based on its well-documented role in monocyte recruitment and the clearance of pneumococcal carriage ^21^. RANTES (CCL5) and IFN-γ were selected due to their elevated levels in carriage-positive individuals and their known involvement in T cell-mediated immune responses ^33^.

We used the data obtained from the initial IL-17A screening of susceptible and protected profiles (Supplementary Figure 3), together with available data on immunisation with those antigens and protection against carriage in mice (24), to down-select a limited number of antigens (Supplementary Table 2) for PBMC stimulation assays. PBMCs from a further 22 carriage-negative and 21 Spn6B carriage-positive participants were stimulated with the down-selected 21 antigens and IL-17A, RANTES, MCP-1 and IFN-γ responses were measured.

### Baseline cytokine/chemokine levels in response to protein antigens are not associate with protection against experimental pneumococcal carriage acquisition with Spn6B

After correction for multiple comparisons, there was no significant difference between baseline protein-specific cytokine/chemokine responses in participants susceptible to experimental colonisation with Spn6B and those who were protected against colonisation (Figure 3). Responses with p-values ≤ 0.05 for the carriage-positive and carriage-negative groups comparison prior to Benjamini-Hochberg correction are shown (Figure 3).

**Figure 3.**
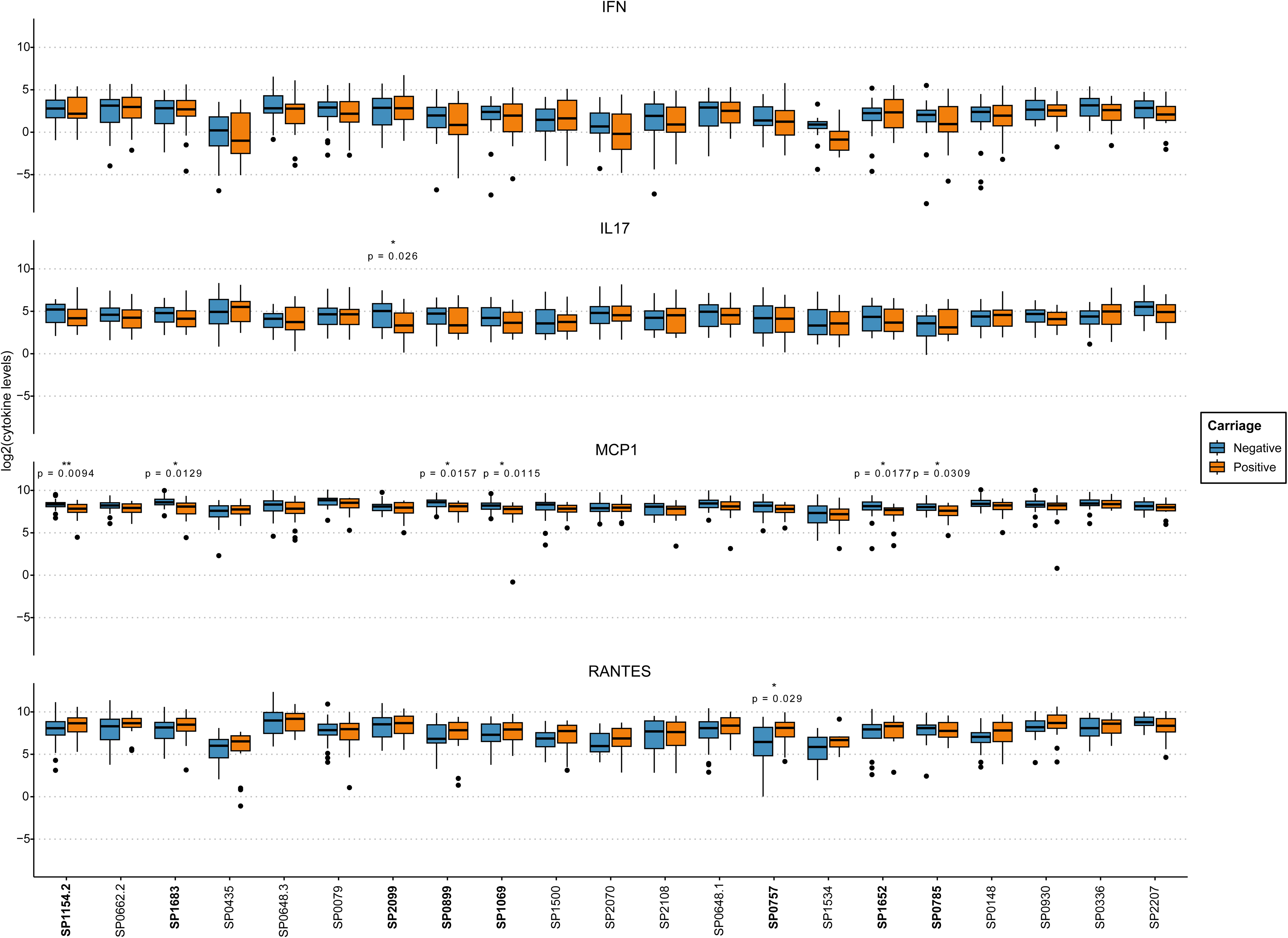
**Cytokine/chemokine responses to pneumococcal proteins**. Baseline levels of cytokines/chemokines produced by PBMCs stimulated with pneumococcal proteins in healthy adults aged 18-50. Bars indicate interquartile range. Spn6B carriage-negative (blue) n = 32, Spn6B carriage-positive (orange) n = 31. Responses with p-value ≤ 0.05 prior to Benjamini-Hochberg correction for multiple comparisons are shown; this significance was lost following correction.

### Random Forest algorithm as an alternative method to rank antigens associated with pneumococcal carriage protection

We used the Random Forest method ^34^ to assess the predictive power of baseline protein-specific antibody (IgG) and cytokine/chemokine levels in predicting susceptibility/protection to carriage acquisition. We trained the model using various features at baseline and selected the combination of features that generated the model with the least error. Antibody response patterns to the proteins PdB, SP0899, and SP1069 together emerged as the most important features, with the model achieving an out-of-bag (OOB) error rate of 36.05%. In terms of cytokine/chemokine responses, IL-17A production in response to SP0648-3 and MCP-1 production in response to SP1069 and SP0899 together were identified as the most relevant features, resulting in an OOB error of 38.98%.

Notably, SP0899 and SP1069 were a feature of both humoral and cellular responses that were predictive of protection against colonisation (Figure 4). Higher baseline IgG against SP1069 and SP0899 was observed in the cohort of participants protected against experimental carriage acquisition (Figure 4). We also observed that those participants protected against colonisation had higher levels of MCP-1 produced in response to stimulation with SP1069 (*p =* 0.035) and SP0899 (*p =* 0.049) than those susceptible to colonisation at baseline (Figure 4).

**Figure 4.**
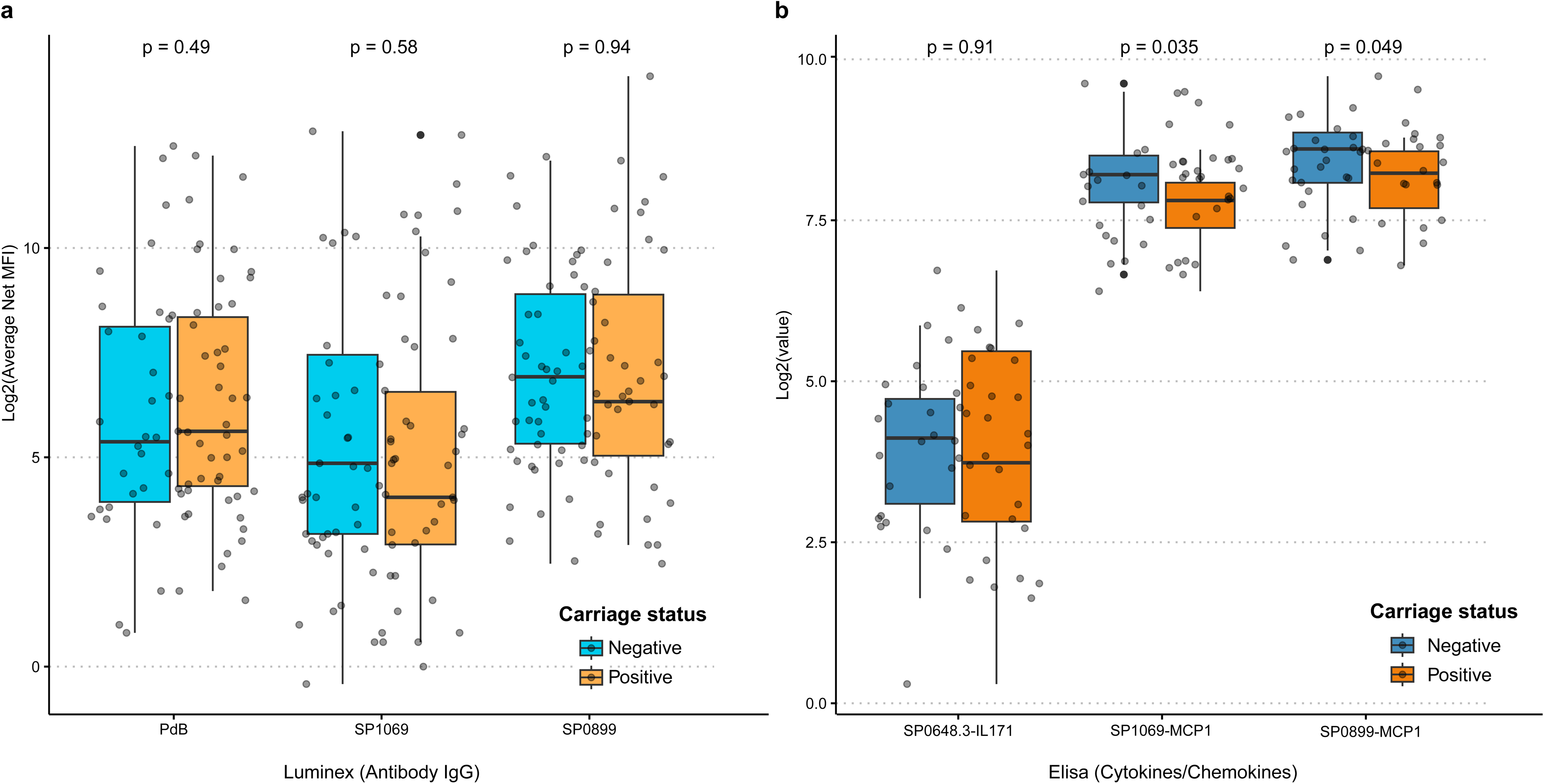
Humoral and cellular responses to the machine learning best ranked proteins antigens. Boxplots showing **(A)** Log2 transformed average net MFI for Spn carriage-positive (orange; n = 36) and Spn carriage-negative (blue; n = 50) participants and **(B)** Log2 transformed cytokine/chemokine concentration for Spn carriage-positive (orange; n = 31) and Spn carriage-negative (blue; n = 32) participants. Each dot represents data from an individual participant. 71

### Correlation of antibody and cytokine/chemokine levels against the proteins selected by machine learning analysis

To determine whether there was a relationship between antibody- and cell-mediated immune responses, we correlated responses identified as predictive of colonisation status by machine learning. Whilst SP0648-3 was found to be correlated in carriage positive participants across datasets (*Rho =* 0.614; *p =* 0.009), SP1069 (*Rho =* 0.378; *p =* 0.203) and SP0899 (*Rho =* −0.242; *p =* 0.425) were not. None of the proteins were correlated in carriage negative participants across datasets (SP0648-3 (*Rho =* −0.076; *p =* 0.781), SP1069 (*Rho =* −0.422; *p =* 0.133), SP0899 (*Rho =* −0.098; *p =* 0.781)).

### Correlation between protein-responses and AUC

We generated area under the curve data for colonising density of the participants colonised with pneumococcus (Figure 5). No significant correlations were found following Benjamini Hochberg correction. Correlations showing *p*-value ≤ 0.05 prior to correction are highlighted in Figure 5 and Supplementary Tables 3 and 4. Participants were stratified by colonisation density and duration, expressed as AUC (Supplementary Figure 4). With respect to cell-mediated immunity, the levels of RANTES, MCP-1, and IFN-γ produced in response to protein stimuli (SP1154-2, SP0648-3, SP0079, and SP1683) differed across density groups (Supplementary Figure 4 B).

**Figure 5.**
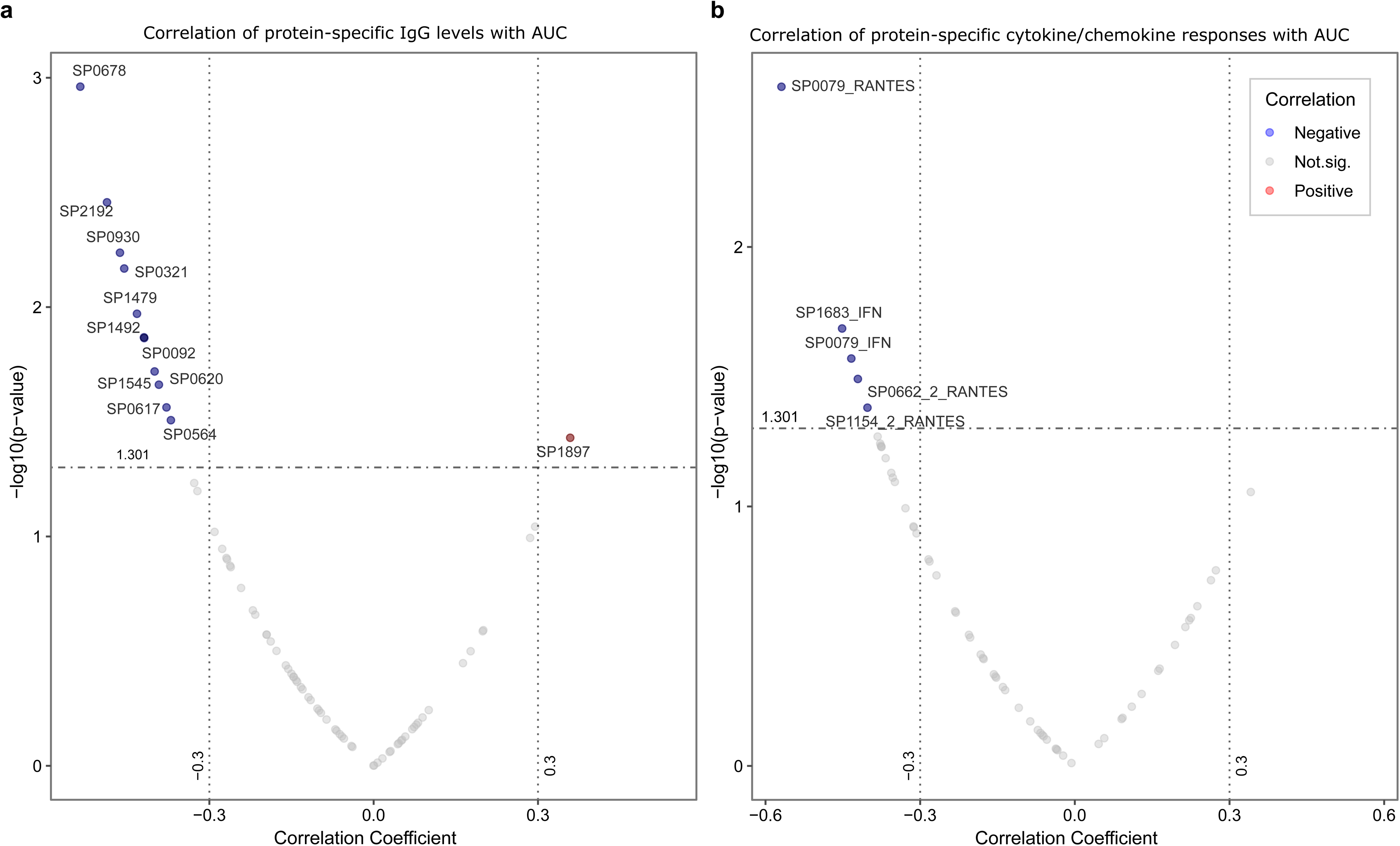
Correlation of protein-specific IgG levels and cytokine/chemokine responses with colonising density (AUC). Correlation results obtained between **(A)** antibody (n= 34) and **(B)** cytokine/chemokine (n= 27) levels for each protein and the AUC values obtained at the interval between days 2 to 14 post-challenge for each Spn6B carriage-positive participant. Proteins with *p*-value ≤0.05 (denoted by horizontal dashed line) were highlighted, blue dots represent protein responses negatively correlated with AUC, while red dots represent the positive correlations. Following Benjamini Hochberg correction for multiple comparisons, all significance was lost.

Although differences were not statistically significant, we observed a trend in high levels of antibodies and cytokines/chemokines specific to protein antigens that were associated with lower colonisation density (Figure 6). We observed that IgG specific to 32 proteins (SP0321, SP0564, SP0346, SP1545, SP1032, SP0617, SP0620, SP0678, SP1479, SP1492, SP0079, SP0402, SP0629, SP0757, SP2145, SP0662-1, SP1683, SP0878, SP0582, SP1376, SP0724, SP1500, SP2083, SP0098, SP0127, SP0604, SP0648-1, SP2207, SP0742, SP0314, SP0648-2, PspA1) was elevated in participants who were either protected against experimental carriage acquisition, or in those who were experimentally colonised at a relatively low bacterial density (Figure 6A). We also observed that cytokine and chemokine responses were elevated in participants who were either protected against experimental carriage acquisition or in the low AUC density group, followed by the mid AUC density and high AUC density groups – IFN-γ and RANTES responses were particularly elevated (Figure 6B). Cytokine and chemokine levels in carriage-negative participants were generally not higher than those observed in the low AUC group. From the selected proteins, only IFN-γ and RANTES responses to SP1154-2 (IFN-γ *p =* 0.034; RANTES *p =* 0.024) and SP0079 (IFN-γ *p =* 0.012; RANTES *p =* 0.005) and the RANTES response to SP0648-1 (p = 0.027) have shown a *p* ≤ 0.05 before Benjamini-Hochberg correction (Figure 6B).

**Figure 6.**
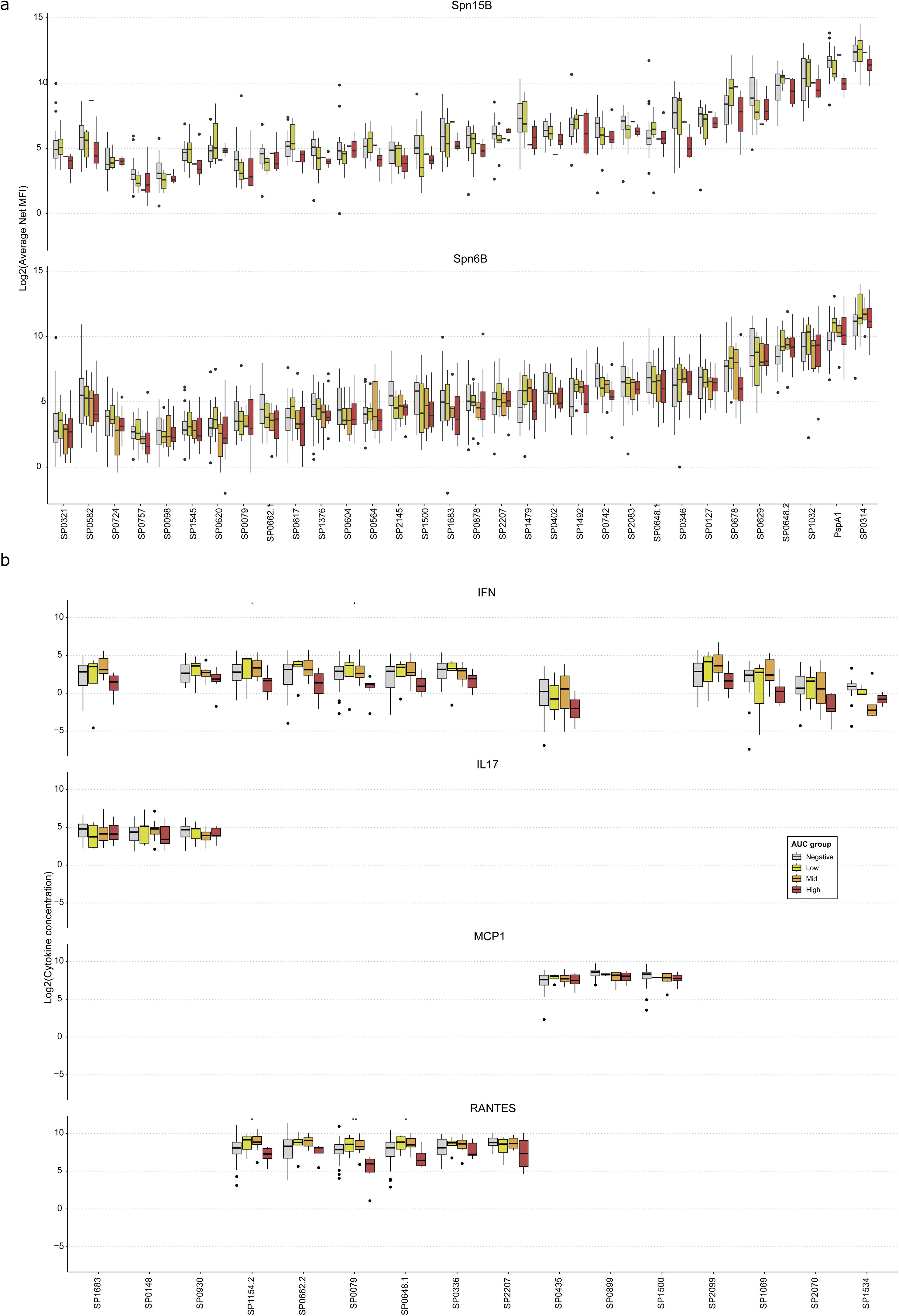
**Humoral and cellular responses across AUC groups for selected proteins showing trend-specific patterns**. **(A)** Baseline levels of anti-protein serum IgG in healthy adults aged 18-50, measured as average net median fluorescence intensity (MFI). Bars indicate interquartile range. Spn15B carriage-negative n = 15, Spn15B carriage-positive n = 13, low AUC n= 8, mid n= 1 and high n= 4; Spn6B carriage-negative n = 18, Spn6B carriage-positive n = 21, low AUC group n= 4, mid n= 6 and high n= 11. **(B)** Baseline levels of selected cytokines/chemokines stimulated by pneumococcal proteins in PBMCs from healthy adults aged 18-50. Carriage-negative - grey; low AUC - yellow, mid AUC - orange; high AUC - red. Bars indicate interquartile range. Spn6B carriage-negative n = 18, Spn6B carriage-positive n = 27, low AUC group n= 4, mid n= 14 and high n= 9.

As previously stated, SP1069 and SP0899 were selected by machine learning for carriage status classification. The IFN-γ response to SP1069 followed the same pattern, levels were highest in the carriage-negative group or in participants colonised at low bacterial density, and lowest in those with high-density colonisation. There was a very marginal difference in the levels of MCP-1 produced in response to SP0899 across groups.

## Discussion

Machine learning and systems biology are increasingly employed in vaccine development to identify and design vaccine candidates as well as to identify correlates of successful immunisation, predict individual vaccine responses, and correlates of protection when a disease/infection endpoint is possible ^35,36^. As the number of measurable biological parameters expands, traditional univariate analyses are inappropriate, increasing the risk of type II errors (failing to detect true associations). In contrast, machine learning enables a more comprehensive and rigorous analysis of complex, high-dimensional datasets, offering deeper insights into the multifactorial nature of immune responses. In this study, we leveraged machine learning to identify immune features predictive of protection against nasal colonisation. Our findings highlight the Random Forest algorithm’s utility in identifying proteins linked to protection against experimental pneumococcal carriage, demonstrating its potential to accelerate vaccine development.

The ultimate aim of pneumococcal vaccination may not be sterilising immunity and complete elimination of nasopharyngeal colonisation, a natural immunising event ^33,36,37,52^, but rather reducing colonisation density to prevent disease progression or transmission to others in the population, particularly during viral co-infections ^53^. Suppressing bacterial density through protein-based vaccines could minimise transmission and reinforce immunity while maintaining mucosal exposure necessary for lung immunity ^54^.

Consistent with prior studies ^36–38^, systemic anti-protein IgG at baseline did not correlate with protection against experimental colonisation by serotype 6B or 15B pneumococcus. Similarly, baseline cell-mediated responses in blood were not predictive of protection in our human cohort, despite evidence of Th17-mediated immunity against pneumococcal colonisation in murine models ^26^. Using machine learning, we identified a group of three antigens — SP1069, SP0899, and PdB — eliciting IgG responses, and a group three antigens — SP1069, SP0899, and SP0648-3 — which stimulated protein-specific cellular responses, that were predictive of protection. Although we cannot yet determine whether these protein combinations could form the basis of a combination vaccine, this possibility warrants further investigation. Notably, SP1069 and SP0899 elicited both humoral and cellular responses. Although the exact functions of these proteins are not yet fully understood, immunisation of mice with a combination of cholera toxin alongside SP1069, SP0899 and SP0648-3 as individual antigens has led to a significant reduction in pneumococcal colonisation compared with the control groups ^26^. Co-administration of multiple antigens with a cholera toxin adjuvant has been shown to enhance the protective efficacy in murine models ^26^. Protein-based vaccines incorporating several antigens may thus offer broad coverage, while conjugating protein antigens to capsular polysaccharides in new PCV formulations could provide additional serotype-independent protection ^39,40^ as well as the advantage of cellular immunity induction.

In humans, protection against pneumococcal disease requires both humoral and cellular immunity ^31,41–46^. Similarly, murine models indicate the necessity of both immunity types for nasopharyngeal colonisation defence ^26,41–43,47,48^. Using our human challenge model, we previously reported that colonisation-induced MCP-1 (CCL2) levels are associated with monocyte recruitment to the nasal mucosa and subsequent clearance of colonisation ^21^. These results also support previous findings from mouse models showing that CCL2 signalling and monocyte recruitment are key mediators of pneumococcal carriage clearance ^49^. Elevated MCP-1 levels in protected participants against experimental acquisition of Spn6B and its involvement in monocyte and neutrophil recruitment underline its significance in pneumococcal clearance^21,50,51^.

In our cohort, baseline anti-protein IgG and cytokine responses correlated with lower colonisation density, suggesting roles for both humoral and cellular immunity in controlling carriage and preventing disease. Consistent with prior findings ^55–58^, IFN-γ and RANTES secretion correlated with reduced colonisation density, likely enhancing innate immune cell recruitment and activity ^59–61^. These results emphasise the importance of designing protein vaccines that elicit both antibody and cellular responses for effective protection.

While promising, this work has limitations. Out-of-bag error rates (∼36–39%) indicate only moderate predictive power, suggesting that further optimisation, such as increasing cohort size or incorporating mucosal immune parameters, may be needed to improve model accuracy. Immune responses to protein antigens in systemic circulation may differ from those in the nasal mucosa, and the study’s small sample size, comprised mainly of healthy young adults, may not reflect high-risk demographics. Validating these findings in samples from older adults participants will be important ^37,62^. Immune correlates may vary across populations with differing pneumococcal burden. Our studies in Malawi ^10,63^ will enable validation across diverse populations and environmental contexts. Further investigation into antigen-specific nasal T-cell responses and sex-stratified immune differences could provide additional insights ^27^.

As immunology and vaccinology increasingly rely on complex, high-dimensional datasets, predictive computational models offer substantial advantages over traditional analyses by uncovering intricate relationships between immune responses and protection. The integration of advanced computational approaches with human challenge models, which uniquely allow sampling immediately before and after defined pathogen exposure, promises to transform how we define protective immunity and accelerate rational vaccine design.

Despite the remarkable success of vaccines, which are estimated to have saved more than 150 million lives over the past 50 years^64^, major challenges persist. Unequal access to vaccines and the biological complexity of difficult-to-target pathogens threaten to stall progress in reducing the global burden of infectious diseases. A key scientific challenge is the identification of causal correlates of protection for high-frequency colonising pathogens such as *Streptococcus pneumoniae*, *Staphylococcus aureus*, and *Escherichia coli*, where repeated antigen exposures at mucosal surfaces shape layered humoral and cellular immune responses. Addressing these gaps will require integrating advanced computational approaches, experimental human models, and global public health strategies to translate immunological insights into next-generation vaccines.

## Data Availability

Data are available upon reasonable request by email directed to the corresponding authors at

## Author Contributions

Conceived the study - DMF, SJ, CS, BU, RM

Data acquisition - KSC, CS, RT, JR, ELG, EM, YL, SP, ENM

Generated the ELISA data - CS

Performed the machine learning analysis - PGD

Performed data analysis - KSC, PGD, CS

Scientific discussion – AJP, SBG, HIN, SHP, ANAG, AMC, SPJ

Project definition and discussion - KSC, PGD, CS, DMF, BU

Provided protein library - RM, YJL, EM, RT

PGD, KSC and CS jointly interpreted the results and wrote the manuscript with contributions from all co-authors. All authors read and approved the final manuscript.

## Acknowledgments

The authors thank all participants in the EHPC studies, as well as the Clinical Research Network North West Coast and the NIHR Comprehensive Local Research Network North West Coast for their continued support. We also extend our gratitude to the study clinical team members for their assistance in sample collection. This work by funded by grants from the Bill & Melinda Gates Foundation (OPP1117728) (to DMF), Medical Research Council (MR/M011569/1) (to SBG and DMF), Robert Austrian Award (to SPJ) and the Bacterial Vaccines (BactiVac) Network funded by the GCRF Networks in Vaccines Research and Development which was co-funded by the MRC and BBSRC (to DMF and CS). KSC was supported by the MRC Doctoral Training Programme (Award 1964515).

