## Supplementary material for "Machine Learning-Driven Identification of Serotype-Independent Pneumococcal Vaccine Candidates using samples from Human Infection Challenge Studies"

^5^BioGrad Limited

^6^Laboratório de Bacteriologia, Instituto Butantan, São Paulo, Brazil

^7^Liverpool University Foundation Hospitals Trust

^8^Department of Clinical and Toxicological Analyses, School of Pharmaceutical Sciences, University of São Paulo, São Paulo, Brazil

^9^Hospital Israelita Albert Einstein, São Paulo, Brazil

^10^Centre for Inflammation Research, University of Edinburgh, Edinburgh, UK.

^11^Boston Children's Hospital, Harvard Medical School, MA, USA

^12^Leiden University Center for Infectious Diseases (LUCID), Leiden University Medical Center, Leiden, Netherlands

**Supplementary material:**

**Supplementary Table 1.** Library of 75 highly conserved pneumococcal proteins used to develop a multiplex assay utilising Luminex xMAP technology.

**Supplementary Text 1.** Luminex assay methodology.

**Supplementary Figure 1.** AUC threshold determination based on retrospective EHPC data

**Supplementary Figure 2.** Carriage-associated cellular responses from supernatants of Spn6B stimulated human PBMCs.

**Supplementary Figure 3.** IL-17A secretion following ex-vivo protein stimulation for all 75 proteins.

**Supplementary Table 2.** The initial IL-17A screening, the data obtained from the susceptible and protected profile together with available data on immunisation with those antigens and protection against carriage in mice was used to down-select a limited number of antigens.

**Supplementary Table 3.** Correlation of protein-specific IgG concentration with colonising density (AUC).

**Supplementary Table 4.** Correlation of cytokine/chemokine responses with colonising density (AUC).

**Supplementary Figure 4.** Humoral and cellular responses across AUC groups for all proteins.

**Supplementary Table 1.** Library of 75 highly conserved pneumococcal proteins used to develop a multiplex assay utilising Luminex xMAP technology.

| **Protein** | **Function** |  | **Protein** | **Function** |
| --- | --- | --- | --- | --- |
| SP0043 | Competence factor transport protein ComB |  | SP1386 | Spermidine/putrescine ABC transporter |
| SP0079* | Potassium uptake protein, Trk family protein |  | SP1404 | Hypothetical protein |
| SP0084 | Sensor histidine kinase |  | SP1479 | Peptidoglycan N-acetylglucosamine deacetylase A |
| SP0092 | ABC^[1]^ transporter, substrate-binding protein |  | SP1500* | Amino acid ABC^1^ transporter substrate-binding protein |
| SP0098 | Hypothetical protein |  | SP1545 | Hypothetical protein |
| SP0127 | Hypothetical protein |  | SP1560 | YbbR-like lipoprotein, putative |
| SP0149 | Lipoprotein |  | SP1652* | Efflux ABC transporter, permease protein |
| SP0191 | Lipoprotein, putative |  | SP1683* | Carbohydrate ABC transporter substrate-binding protein |
| SP0198 | Hypothetical protein |  | SP1826 | ABC transporter, substrate-binding protein |
| SP0249 | PTS^[2]^ system, IIB component |  | SP1872 | Iron-compound ABC transporter, iron-compound-binding protein |
| SP0321 | PTS system, IIA component |  | SP1897 | Sugar ABC transporter, sugar-binding protein |
| SP0346 | Capsular  polysaccharide  biosynthesis protein Cps4A |  | SP1942 | Transcriptional regulator, putative |
| SP0402 | Signal peptidase I |  | SP2083 | Sensor histidine kinase PnpS |
| SP0435* | Translation elongation factor P |  | SP2151 | Carbamate kinase |
| SP0564 | Hypothetical protein |  | SP2192 | Sensor histidine kinase |
| SP0582 | Hypothetical protein |  | SP2197 | ABC transporter, substrate-binding protein, putative |
| SP0601 | ABC transporter, transmembrane protein Vexp3 |  | SP2207* | Competence protein ComF, putative |
| SP0604 | Sensor histidine kinase VncS |  | SP2218 | Rod shape-determining protein MreC |
| SP0617 | Matrixin family protein |  | SP2145 | Cell wall surface anchor protein |
| SP0620 | Amino acid ABC transporter, amino acid-binding protein, putative |  | SP1534* | Manganese-dependent inorganic pyrophosphatase, putative |
| SP0629 | D-Ala-D-Ala carboxypeptidase, metallo peptidase |  | SP2070* | Glucose-6-phosphate isomerase |
| SP0648-1* | Beta-galactosidase |  | SP1002 | Adhesion lipoprotein |
| SP0648-2 | Beta-galactosidase |  | SP0148* | ABC transporter, substrate-binding protein |
| SP0648-3* | Beta-galactosidase |  | SP0314 | Hyaluronate lyase precursor |
| SP0659 | Thioredoxin family protein |  | SP0336* | Penicillin-binding protein |
| SP0662-2* | Sensor histidine kinase |  | SP0369 | Penicillin-binding protein 1A |
| SP0678 | Rhodanese-like domain |  | SP0930* | Choline binding protein E |
| SP0724 | Hypothetical protein |  | SP1492 | Cell wall surface anchor family protein |
| SP0742 | Hypothetical protein |  | SP1833 | Cell wall surface anchor family protein, metal binding |
| SP0757* | Cell division protein FtsX |  | SP2108* | Maltose/maltodextrin ABC transporter, maltose/maltodextrin-binding protein |
| SP0785* | HlyD family secretion protein |  | SP0662-1 | Sensor histidine kinase, putative |
| SP0787 | Antimicrobial peptide ABC transporter permease |  | SP2099* | Penicillin-binding protein 1B |
| SP0878 | SpoE family protein |  | PspC6 | Pneumococcal surface protein C (clade 6); binds complement factor H to evade C3 opsonization |
| SP0899* | Hypothetical protein |  | PspC9 | Pneumococcal surface protein C (clade 9); binds complement factor H to evade C3 opsonization |
| SP1032 | Iron-compound ABC transporter |  | PspA1 | Pneumococcal surface protein A (clade 1); cell wall-associated protein involved in inhibiting complement-mediated opsonization and preventing lactoferrin-mediated clearance |
| SP1069* | Hypothetical protein |  | PspA4 | Pneumococcal surface protein A (clade 4); cell wall-associated protein involved in inhibiting complement-mediated opsonization and preventing lactoferrin-mediated clearance |
| SP1154-2* | IgA1-specific metallopeptidase |  | PdB | Pneumolysin mutant |
| SP1376 | Shikimate 5-dehydrogenase |  |  | |

^[1]^ ATP-binding cassette

^[2]^ Phosphotransferase system

**Supplementary Text 1. Luminex assay methodology**

Proteins were randomly allocated to different microsphere regions. As 22 regions were available, proteins were divided across 4 panels. A ‘master-mix’ of each panel was made containing 250 beads/µl of each protein-coupled bead population (stored in wash buffer). These stocks were protected from light and stored at 4°C. From this stock solution, the required volume of stock microspheres was removed and diluted 1:5 in 1%BSA-PBS (assay buffer) to achieve 50 beads/µl of each protein-coupled bead population. 25µl of bead solution was added to each well on a 96 well plate. To this, 25µl of 1:20,000 serum diluted in an assay buffer was added. A stock of pooled sera taken 29 days post-challenge and diluted 1:50 in assay buffer was used as an internal positive control across all plates and assay buffer was used as a negative control to detect background median fluorescence intensity (MFI) values. The plate was covered to protect microspheres from light and incubated overnight at 4°C on a plate shaker (300rpm). Following incubation, the plate was clipped into place on the Luminex Magnetic Plate Separator and rapidly and forcefully inverted over a biohazard receptacle to expel the liquid from the wells. Microspheres were washed three times with 200µl assay buffer. Biotinylated human IgG Fc antibody (Invitrogen™ Novex™, catalogue number: A18821, lot: 62, 129, 090718) was diluted to 2µg/ml in assay buffer and 50µl was then added to each well. The plate was then incubated in the dark for 30 minutes at room temperature on a plate shaker. Following incubation, liquid was expelled from wells and microspheres were washed three times as previously. Streptavidin R-phycoerythrin conjugate (SA-PE) (Invitrogen, catalogue number: S866, lot: 1973501) was diluted idem as for the biotinylated secondary antibody and 50µl of SA-PE solution was added to each well. The plate was incubated as previously for 10 minutes. Liquid was expelled from wells and microspheres were then washed three times and re-suspended in 50µl of assay buffer. The plate was incubated for five minutes before analysing the contents of each well on the Luminex analyser. All samples were run in duplicate. Serum samples from two participant cohorts, as described above, were analysed. All data reported are in units of average net MFI (average of replicates) with background MFI subtracted (average net MFI - blank wells). It should be noted that the internal positive control both confirmed immunodetection as well as inter-assay variation of the panel of proteins against which serum IgG was being analysed. In the case of high variation between plates, experiments were repeated to ensure comparability of results (<25% CV for each analyte). Final inter-assay CV for all proteins as determined by the internal positive control was <20%. Thus, we deem results obtained across different assays to be comparable.

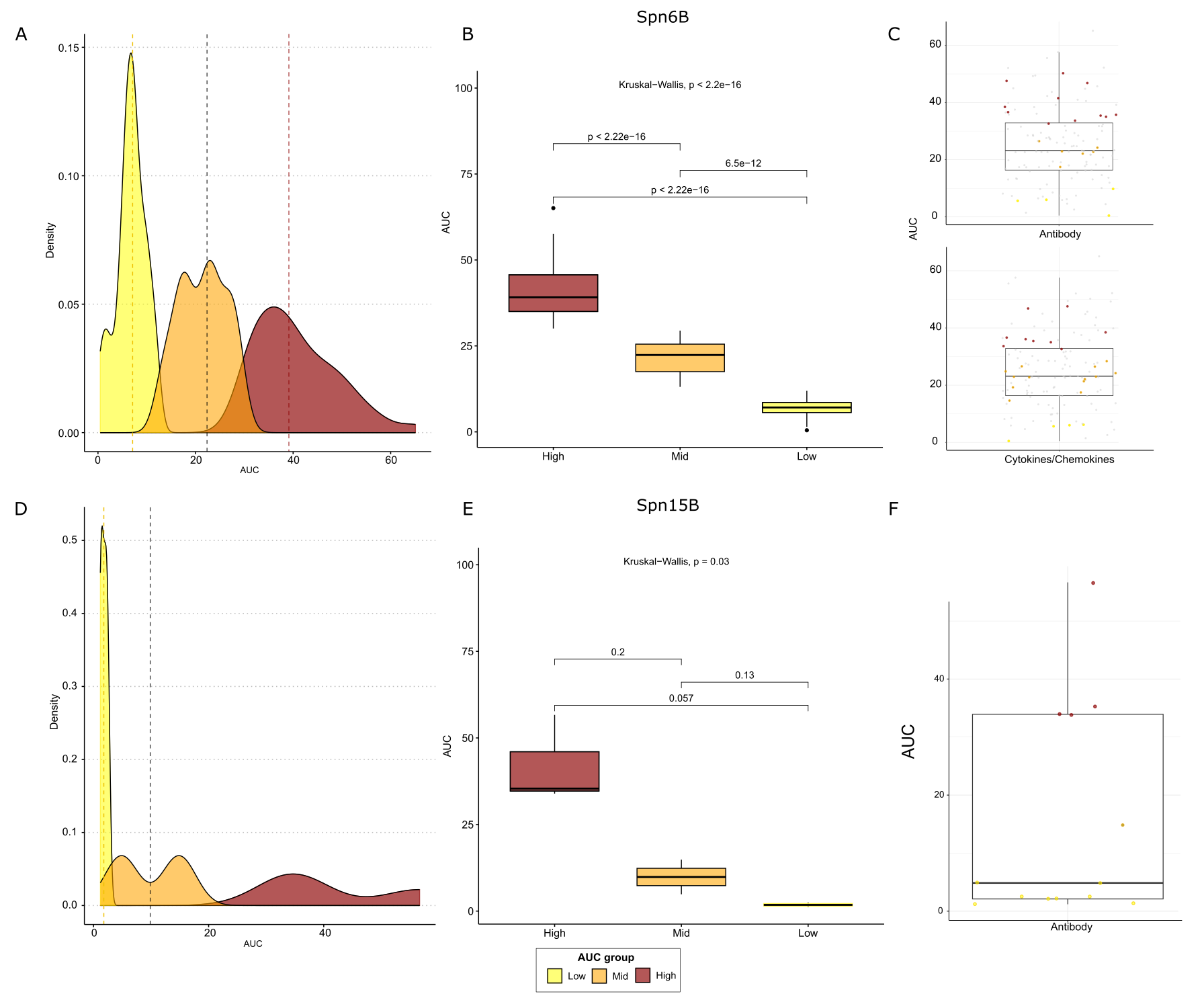

**Supplementary Figure 1. AUC threshold determination based on retrospective EHPC data. (A)** Density plot of AUC values categorized into three groups: Low (yellow), Mid (orange), and High (dark red). Vertical dashed lines indicate threshold values separating these groups. **(B)** Box plot comparing AUC distributions across Low, Mid, and High groups for Spn6B. Kruskal-Wallis p-value and pairwise comparisons are shown. **(C)** Box plots displaying AUC distributions for Antibody and Cytokines/Chemokines responses. Grey dots represent AUC values from retrospective studies, while coloured dots represent AUC values from Spn15B participants in this study. Participant data are categorized as follows: yellow (Low, below Q1), orange (Mid, Q1-Q3), and dark red (High, above Q3). **(D)** Density plot of AUC values for Spn15B, categorized into Low, Mid, and High groups, similar to (A). **(E)** Box plot comparing AUC distributions for Spn15B across Low, Mid, and High groups. Kruskal-Wallis p-value and pairwise comparisons are shown. **(F)** Box plot displaying AUC distribution for the Antibody response. Grey dots represent AUC values from retrospective studies, while coloured dots represent AUC values from Spn15B participants in this study. Participant data are categorised as follows: yellow (Low, below Q2), orange (Mid, Q2), and dark red (High, above Q3).

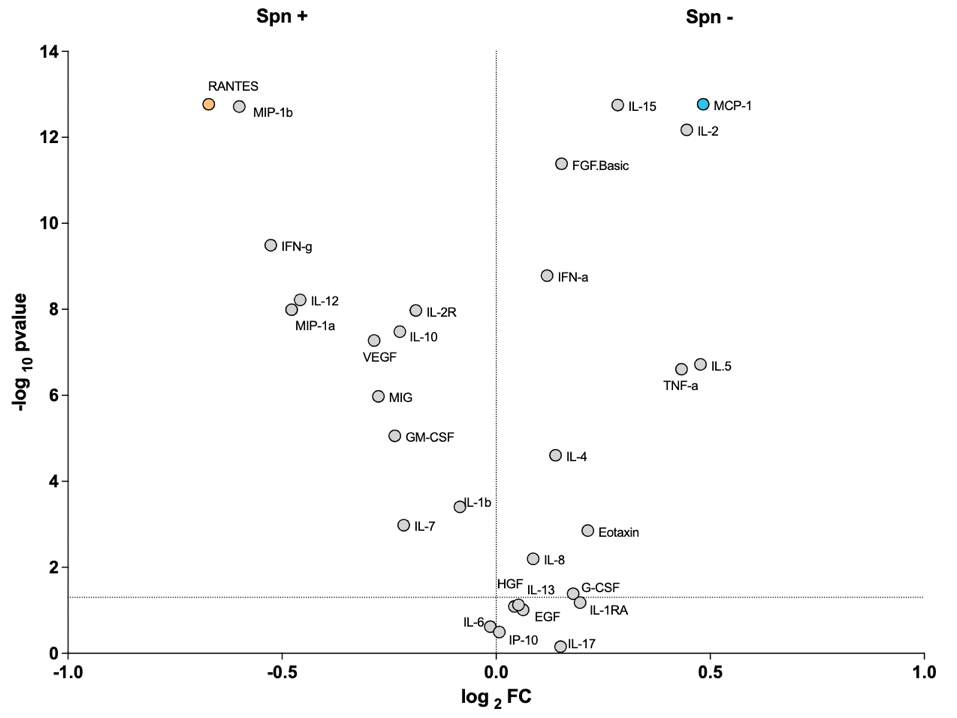

**Supplementary Figure 2. Carriage-associated cellular responses from supernatants of Spn6B stimulated human PBMCs.** Supernatants from PBMC cultures stimulated for 7 days with a protein library and heat-inactivated Streptococcus pneumoniae serotype 6B (Spn6B) were analyzed for cytokine and chemokine concentrations using a 30-plex human Luminex magnetic bead assay. Samples were derived from a pooled set of 12 carriage-negative (Spn–) and 11 carriage-positive (Spn+) healthy adult participants. Each dot represents an individual analyte, plotted by its log_2_(fold change) (Spn– vs Spn+) on the x-axis and the –log_10_(p-value) (Wilcoxon paired test) on the y-axis. The horizontal dashed line indicates the p = 0.05 threshold. Analytes to the right of the vertical axis (positive log_2_FC) are elevated in the Spn– group, while those to the left are elevated in the Spn+ group.

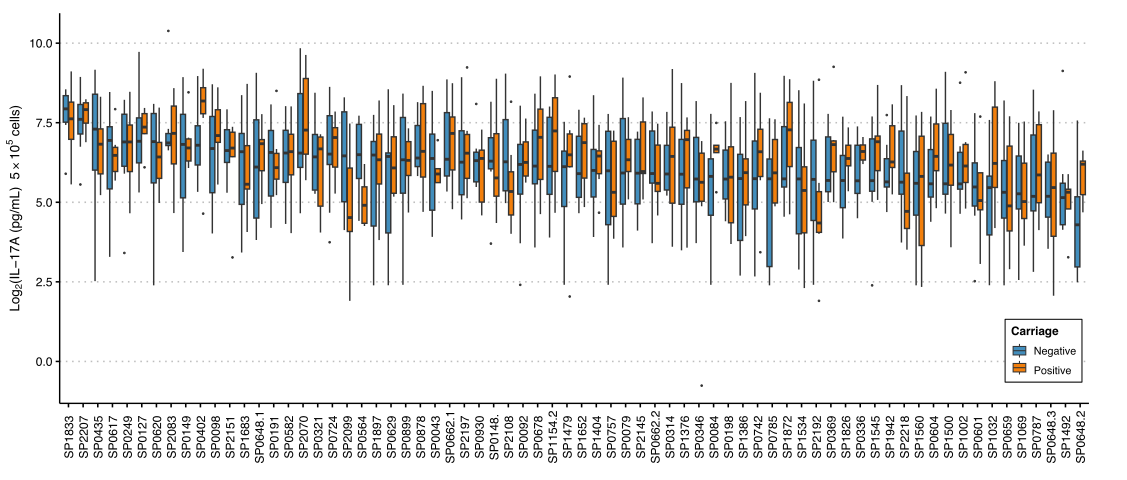

**Supplementary Figure 3.** **IL-17A secretion following ex-vivo protein stimulation for all 75 proteins.** Baseline levels of IL-17 produced by PBMCs stimulated with pneumococcal proteins in healthy adults aged 18-50. Spn6B carriage-negative (blue) n = 10, Spn6B carriage-positive (orange) n = 11.

**Supplementary Table 2.** The initial IL-17A screening, the data obtained from the susceptible and protected profile together with available data on immunisation with those antigens and protection against carriage in mice was used to down-select a limited number of antigens.

|  |  | **Correlations** | | | **High Neg >FC 0.5** | **High Pos <FC -0.5** | |
| --- | --- | --- | --- | --- | --- | --- | --- |
| **Gene name** | **Mice Data** | **D2** | **D7** | **AUC D2-D7** | **MCP-1** | **IFN-g** | **RANTES** |
| ***SP0079*** |  |  | -0.78 |  | 0.533 | -0.507 |  |
| ***SP0148*** | YES |  |  |  |  | -1.805 | -1.204 |
| ***SP0336**** |  |  |  |  |  |  | -0.575 |
| ***SP0435*** | YES | -1 |  |  | 0.480 |  | -0.930 |
| ***SP0648-1*** | YES |  |  |  | 0.680 |  | -0.829 |
| ***SP0648-3*** |  | -0.88 |  |  | 0.708 | -0.483 | -0.765 |
| ***SP0662-2*** | YES |  | -0.84 |  | 0.491 | -1.125 | -1.343 |
| ***SP0757*** | YES | no sig -0.82 |  |  |  | -1.449 | -1.020 |
| ***SP0785*** | YES | no sig -0.82 |  | -1 |  | -2.263 | -0.588 |
| ***SP0899*** | YES |  |  |  |  | -2.563 | -0.880 |
| ***SP0930**** |  |  |  |  |  |  |  |
| ***SP1069*** | YES |  |  |  |  | -2.451 | -0.857 |
| ***SP1154-2*** | YES | -1 |  | no sig -0.82 |  | -1.485 | -0.715 |
| ***SP1500*** | YES |  |  |  |  | -2.418 | -0.901 |
| ***SP1534*** | YES |  |  | -0.94 |  |  |  |
| ***SP1652*** |  |  |  | -0.88 |  | -2.082 | -0.557 |
| ***SP1683*** | YES |  | -0.88 |  |  | -1.526 | -0.665 |
| ***SP2070*** | YES |  |  |  | 0.496 |  | -0.735 |
| ***SP2099*** |  |  | -0.78 | no sig -0.75 | 0.508 |  | -0.893 |
| ***SP2108*** | YES |  |  |  | 0.596 | -2.127 | -0.915 |
| ***SP2207**** |  |  |  |  |  | -0.850 | -0.589 |

*proteins that did not meet the criteria but were included as controls

*c- : carriage-negative, c+: carriage-positive, FC: Fold Change, AUC: Area under the curve

**Supplementary Table 3.** Correlation of protein-specific IgG concentration with colonising density (AUC).

| **IgG levels to:** | **Rho** | **p-value** | **BH adjusted p-value** |
| --- | --- | --- | --- |
| SP0678 | -0.536 | 0.001 | 0.079 |
| SP2192 | -0.487 | 0.003 | 0.122 |
| SP0930 | -0.463 | 0.006 | 0.122 |
| SP0321 | -0.455 | 0.007 | 0.122 |
| SP1479 | -0.432 | 0.011 | 0.140 |
| SP0092 | -0.419 | 0.014 | 0.140 |
| SP1492 | -0.419 | 0.014 | 0.140 |
| SP1545 | -0.400 | 0.019 | 0.172 |
| SP0620 | -0.392 | 0.022 | 0.174 |
| SP0617 | -0.378 | 0.027 | 0.197 |
| SP0564 | -0.370 | 0.031 | 0.204 |
| SP1942 | -0.328 | 0.058 | 0.324 |
| SP2218 | -0.322 | 0.063 | 0.326 |
| SP0659 | -0.290 | 0.096 | 0.430 |
| SP0149 | -0.277 | 0.113 | 0.445 |
| SP2197 | -0.269 | 0.124 | 0.445 |
| SP0198 | -0.268 | 0.126 | 0.445 |
| SP0757 | -0.262 | 0.134 | 0.445 |
| SP0402 | -0.261 | 0.136 | 0.445 |
| SP0582 | -0.242 | 0.168 | 0.525 |
| SP0191 | -0.220 | 0.210 | 0.630 |
| SP0314 | -0.216 | 0.219 | 0.632 |
| SP1683 | -0.195 | 0.268 | 0.665 |
| SP0785 | -0.195 | 0.268 | 0.665 |
| SP1032 | -0.188 | 0.287 | 0.689 |
| PspA1 | -0.177 | 0.315 | 0.712 |
| SP0662.1 | -0.160 | 0.365 | 0.772 |
| SP1872 | -0.156 | 0.378 | 0.775 |
| SP0742 | -0.150 | 0.397 | 0.775 |
| SP0878 | -0.146 | 0.409 | 0.775 |
| SP0346 | -0.146 | 0.410 | 0.775 |
| SP1560 | -0.142 | 0.423 | 0.775 |
| SP2207 | -0.140 | 0.431 | 0.775 |
| SP2099 | -0.133 | 0.454 | 0.795 |
| SP0648.2 | -0.130 | 0.464 | 0.795 |
| SP1404 | -0.119 | 0.502 | 0.841 |
| SP0127 | -0.115 | 0.517 | 0.846 |
| SP1376 | -0.103 | 0.563 | 0.879 |
| SP2083 | -0.100 | 0.574 | 0.879 |
| PspA4 | -0.096 | 0.588 | 0.882 |
| SP1500 | -0.086 | 0.628 | 0.903 |
| SP1534 | -0.070 | 0.694 | 0.903 |
| SP0662.2 | -0.068 | 0.703 | 0.903 |
| SP0724 | -0.062 | 0.727 | 0.903 |
| PspC9 | -0.058 | 0.744 | 0.903 |
| SP2108 | -0.054 | 0.760 | 0.903 |
| SP0648.1 | -0.041 | 0.819 | 0.903 |
| SP1826 | -0.039 | 0.827 | 0.903 |
| PdB | -0.0004 | 0.998 | 0.998 |
| SP0601 | 0.001 | 0.997 | 0.998 |
| SP0899 | 0.007 | 0.968 | 0.996 |
| SP0336 | 0.016 | 0.929 | 0.969 |
| SP0435 | 0.029 | 0.870 | 0.922 |
| SP0369 | 0.031 | 0.862 | 0.922 |
| SP0648.3 | 0.044 | 0.805 | 0.903 |
| SP0604 | 0.046 | 0.797 | 0.903 |
| PspC6 | 0.050 | 0.778 | 0.903 |
| SP1069 | 0.052 | 0.770 | 0.903 |
| SP0249 | 0.056 | 0.746 | 0.903 |
| SP0148 | 0.070 | 0.692 | 0.903 |
| SP0098 | 0.074 | 0.678 | 0.903 |
| SP1154.2 | 0.078 | 0.661 | 0.903 |
| SP0079 | 0.081 | 0.649 | 0.903 |
| SP1386 | 0.089 | 0.615 | 0.903 |
| SP0787 | 0.100 | 0.572 | 0.879 |
| SP0629 | 0.163 | 0.357 | 0.772 |
| SP1002 | 0.177 | 0.317 | 0.712 |
| SP2151 | 0.199 | 0.259 | 0.665 |
| SP0043 | 0.200 | 0.256 | 0.665 |
| SP1833 | 0.286 | 0.101 | 0.430 |
| SP1652 | 0.295 | 0.091 | 0.430 |
| SP1897 | 0.359 | 0.037 | 0.223 |

**Supplementary Table 4.** Correlation of cytokine/chemokine responses with colonising density (AUC).

| **Cytokine/Chemokine Levels to:** | **Rho** | **p-value** | **BH adjusted p-value** |
| --- | --- | --- | --- |
| SP0079_RANTES | -0.569 | 0.002 | 0.147 |
| SP1683_IFN | -0.451 | 0.021 | 0.351 |
| SP0079_IFN | -0.434 | 0.027 | 0.351 |
| SP0662_2_RANTES | -0.421 | 0.032 | 0.351 |
| SP1154_2_RANTES | -0.402 | 0.042 | 0.351 |
| SP2070_IFN | -0.383 | 0.054 | 0.351 |
| SP0662_2_IFN | -0.378 | 0.057 | 0.351 |
| SP2099_RANTES | -0.376 | 0.058 | 0.351 |
| SP0648_1_RANTES | -0.375 | 0.059 | 0.351 |
| SP1154_2_IFN | -0.375 | 0.059 | 0.351 |
| SP0899_RANTES | -0.367 | 0.065 | 0.351 |
| SP1683_RANTES | -0.356 | 0.074 | 0.351 |
| SP0648_3_IFN | -0.353 | 0.077 | 0.351 |
| SP1069_RANTES | -0.349 | 0.080 | 0.351 |
| SP0648_1_IFN | -0.329 | 0.102 | 0.387 |
| SP1500_RANTES | -0.313 | 0.119 | 0.408 |
| SP0757_RANTES | -0.312 | 0.120 | 0.408 |
| SP0899_IFN | -0.307 | 0.127 | 0.408 |
| SP2099_IFN | -0.284 | 0.160 | 0.473 |
| SP1500_IFN | -0.282 | 0.163 | 0.473 |
| SP0648_3_RANTES | -0.268 | 0.184 | 0.489 |
| SP1069_IFN | -0.232 | 0.254 | 0.578 |
| SP0435_IL17 | -0.231 | 0.257 | 0.578 |
| SP0648_1_MCP1 | -0.205 | 0.312 | 0.610 |
| SP2108_RANTES | -0.203 | 0.320 | 0.610 |
| SP0435_IFN | -0.183 | 0.372 | 0.657 |
| SP2070_RANTES | -0.178 | 0.384 | 0.657 |
| SP1069_MCP1 | -0.177 | 0.387 | 0.657 |
| SP1534_IFN | -0.157 | 0.443 | 0.680 |
| SP2108_IFN | -0.154 | 0.452 | 0.680 |
| SP1534_IL17 | -0.153 | 0.457 | 0.680 |
| SP0899_MCP1 | -0.140 | 0.497 | 0.721 |
| SP0435_RANTES | -0.135 | 0.510 | 0.724 |
| SP2070_MCP1 | -0.109 | 0.597 | 0.792 |
| SP0079_IL17 | -0.086 | 0.673 | 0.838 |
| SP0079_MCP1 | -0.072 | 0.727 | 0.878 |
| GenderFemale | -0.067 | 0.746 | 0.878 |
| SP1069_IL17 | -0.061 | 0.769 | 0.878 |
| SP1500_IL17 | -0.054 | 0.792 | 0.878 |
| SP0662_2_MCP1 | -0.037 | 0.858 | 0.899 |
| SP0757_MCP1 | -0.036 | 0.863 | 0.899 |
| SP1500_MCP1 | -0.034 | 0.870 | 0.899 |
| SP1534_RANTES | -0.023 | 0.912 | 0.928 |
| SP0757_IFN | -0.007 | 0.975 | 0.975 |
| SP2070_IL17 | 0.046 | 0.823 | 0.896 |
| SP0435_MCP1 | 0.057 | 0.782 | 0.878 |
| SP0648_1_IL17 | 0.091 | 0.660 | 0.838 |
| SP1683_MCP1 | 0.093 | 0.652 | 0.838 |
| SP1154_2_MCP1 | 0.110 | 0.591 | 0.792 |
| SP2099_IL17 | 0.130 | 0.528 | 0.732 |
| SP0648_3_MCP1 | 0.162 | 0.430 | 0.680 |
| SP2099_MCP1 | 0.165 | 0.422 | 0.680 |
| SP0757_IL17 | 0.194 | 0.342 | 0.632 |
| SP0648_3_IL17 | 0.214 | 0.292 | 0.593 |
| SP1683_IL17 | 0.222 | 0.275 | 0.578 |
| SP1652_IL17 | 0.225 | 0.269 | 0.578 |
| SP0662_2_IL17 | 0.238 | 0.242 | 0.578 |
| SP2108_IL17 | 0.264 | 0.192 | 0.489 |
| SP0899_IL17 | 0.274 | 0.176 | 0.489 |
| SP1154_2_IL17 | 0.341 | 0.088 | 0.358 |

**Supplementary Figure 4. Humoral and cellular responses across AUC groups for all proteins.**

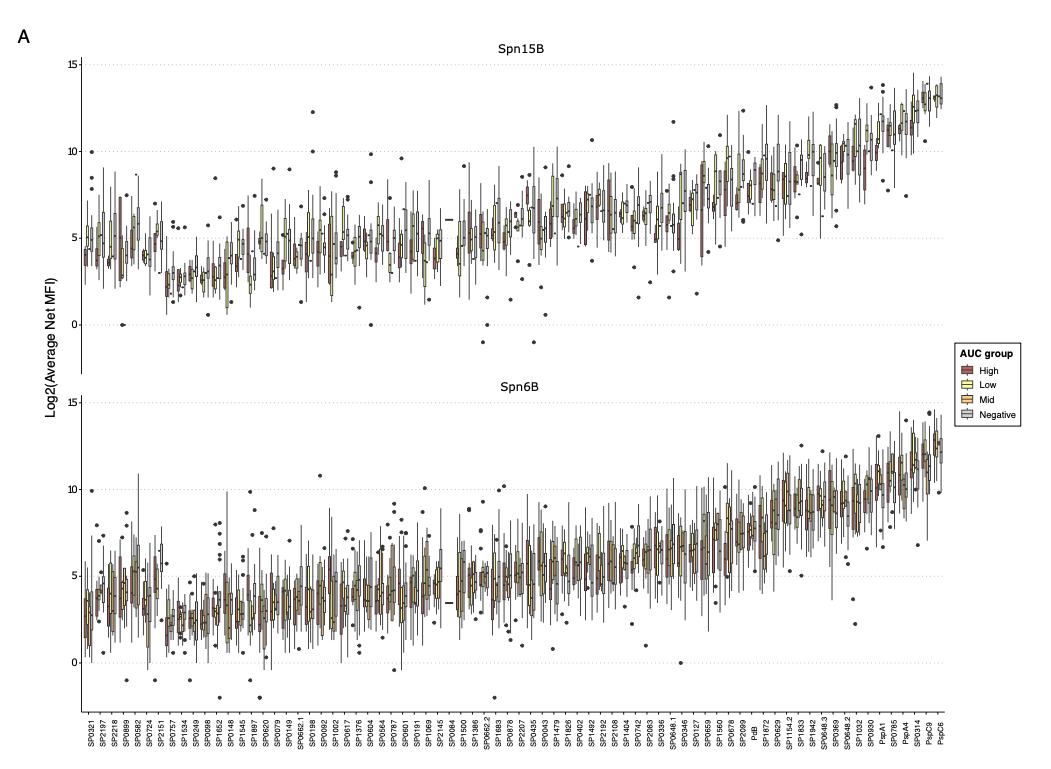

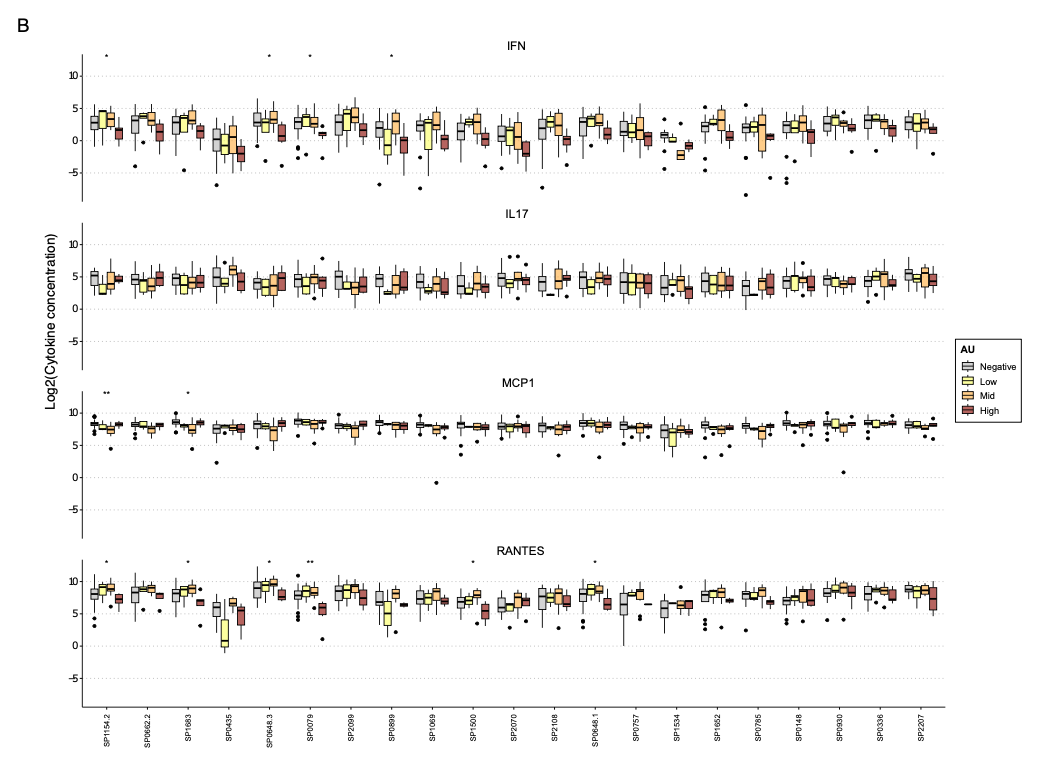

**Supplementary Figure 4. Humoral and cellular responses across AUC groups for all proteins**. **(A)** Baseline levels of anti-protein serum IgG in healthy adults aged 18-50, measured as average net median fluorescence intensity (MFI). Bars indicate interquartile range. Spn15B carriage-negative n = 15, Spn15B carriage-positive n = 13, low AUC n= 8, mid n= 1 and high n= 4; Spn6B carriage-negative n = 18, Spn6B carriage-positive n = 21, low AUC group n= 4, mid n= 6 and high n= 11. **(B)** Baseline levels of selected cytokines/chemokines produced by PBMCs from healthy adults aged 18-50 that were stimulated by pneumococcal proteins. Carriage-negative - grey; low AUC - yellow, mid AUC - orange; high AUC - red. Bars indicate interquartile range. Spn6B carriage-negative n = 18, Spn6B carriage-positive n = 27, low AUC group n= 4, mid n= 14 and high n= 9.
